# Effect of intravesical chemotherapy on the survival of patients with non-muscle-invasive bladder cancer undergoing transurethral resection: a retrospective cohort study among older adults

**DOI:** 10.1101/2021.03.11.21253269

**Authors:** Ashis Kumar Das, Devi Kalyan Mishra, Saji Saraswathy Gopalan

## Abstract

**Background:** The average age of diagnosis for bladder cancer is 73 and about 75 percent of all bladder cancers are non-muscle invasive at initial diagnosis. It is recommended that non-muscle invasive bladder cancers (NMIBC) should be treated with transurethral resection of the bladder tumor (TURBT) followed by chemotherapy. However, there is no large-scale study from real-world databases to show the effectiveness of chemotherapy on the survival of older adults with NMIBC that have undergone TURBT. This study aimed to investigate the effects of chemotherapy on survival among older NMIBC patients with TURBT.

**Methods:** Using the Surveillance, Epidemiology, and End Results (SEER) database (2010-2015), we performed analyses of cancer-specific mortality and overall mortality comparing chemotherapy versus no chemotherapy after TURBT. Coarsened exact matching was performed to balance the baseline patient characteristics. Cox proportional hazards and Kaplan-Meir analyses were used to evaluate survival outcomes.

**Results:** A total of 3,222 matched patients with 1,611 in each arm (chemotherapy and no chemotherapy) were included in our study. After adjusting for covariates, multivariable Cox regression analyses show chemotherapy was associated with lower cancer-specific mortality (HR 0.63; 95% CI 0.42-0.94; p value 0.024). However, chemotherapy did not have any effect on overall mortality (HR 0.84; 95% CI 0.65-1.07; p value 0.159). The Kaplan-Meier curves show the protective effects of chemotherapy on cancer specific survival (p=0.032), but not on overall survival (p=0.34).

**Conclusion:** Chemotherapy improved cancer specific survival among older patients with non-muscle invasive bladder cancer undergoing TURBT surgery, but it had no effect on overall survival. There is a need for more granular level real-world data on chemotherapy regimens and dosage to effectively investigate the effects of chemotherapy on survival of older patients with NMIBC that have undergone TURBT.

## Introduction

Bladder cancer is the most common cancer of the urinary system among the American population, and an estimated 83,730 new cases of bladder cancer and 17,200 deaths are expected to occur in the United States in 2021.^1^ It is the fourth most common cancer among men. The average age of diagnosis for bladder cancer is 73 and about 90 percent of patients are above 55 years.^2^ About 75 percent of all bladder cancers are non-invasive at initial diagnosis.^3^ The non-muscle invasive bladder cancers (NMIBC) include papillary tumors (Ta), carcinoma in situ (CIS or Tis) and tumors invading the subepithelial connective tissue (T1). The 5-year relative survival rate for bladder cancer is 77% overall, whereas it can be up to 96% for patients with NMIBC.^2^ Though patients with NMIBC have higher survival rates, they have a high chance of recurrence or progression even with therapies.^3^ Depending on the clinical and pathological characteristics, NMIBCs have been further risk-stratified as low-, intermediate- and high-risk groups. The recent guidelines recommend that low- and intermediate-risk NMIBC should be treated with transurethral resection of the bladder tumor (TURBT) followed by a single postoperative instillation of intravesical chemotherapy within 24 hours of TURBT.^4^

While the guidelines recommend and clinical trials have shown the benefits, there is no large-scale study from real-world databases to show the effectiveness of chemotherapy on the survival of older adults with NMIBC that have undergone TURBT.^4–7^ Therefore, the aim of this study was to estimate the additional benefit of intervascular chemotherapy on survival of older adults with NMIBC that have undergone TURBT.

## Materials and Methods

### Study population

We selected patients from the Surveillance Epidemiology and End Result (SEER) 21 databases.^8^ SEER database has a completeness rate of 98% and all patients were followed up for 10 years after routine treatment until death or loss to follow-up.^9^ Patient details such as demographic background, tumor features, and survival are available from these databases. We included patients in our study if they met the following criteria: (a) year of diagnosis 2010 to 2015; (b) adults of 65 years or older age; (c) non-muscle invasive bladder cancer (T1, Ta and Tis); (d) positive histology; and (e) undergone transurethral resection of bladder tumor (TURBT). We excluded patients if they had: (a) radiation therapy; (b) missing socio-demographic characteristics – age, sex, race; (c) missing survival time; (d) missing/unknown cause of death classification; (e) missing tumor characteristics – tumor size, number of malignant tumors, (f) tumor with lymph node involvement; (g) high-risk NMIBC as defined by American Urological Association; and (g) metastatic tumor.

### Variables definition

The outcome variables for our study were cancer specific (CSM) and overall mortality (OM). Censoring was applied to patients that died of other causes or were still alive at the end of the follow-up period. Chemotherapy – adjuvant or neo-adjuvant – was coded as either administered or not administered according to the SEER classification. Covariates were patient level demographic and tumor specific variables as well as year of diagnosis. Demographic covariates were sex (male, female), age at diagnosis (categorized by years 65-70, 71-75, 76-80, >80), and race (Black, White, others). Tumor specific covariates were grade (I, II, III, IV and unknown), site (trigone, lateral wall, posterior wall, overlapping lesion of bladder, bladder NOS and others), tumor size (up to 10 mm, 11-20 mm, 21-30 mm, 31-40 mm, 41-50 mm and above 50 mm), and number of in-situ tumors (solitary, multiple).

### Statistical analyses

Frequencies and proportions of categorical variables constituted descriptive statistics. Differences in proportions for categorical variables were compared by the chi-square test. There were baseline imbalances among the covariates. So, a matching technique known as coarsened exact matching was used to create a balanced sample.^10^ The cancer specific survival and overall survival curves were created by the Kaplan-Meier method with the log-rank test to compare survival between groups. Multivariable Cox regressions were performed to estimate the impact of chemotherapy on prognosis. The results of the regressions were presented as hazard ratios (HR) with their 95% confidence intervals (CI). The statistical analyses were performed using R programming language (version 3.6.3) and Stata (version 15). All reported P-values were two-tailed with a level of significance at P < 0.05.

## Results

### Descriptive analysis

Applying the inclusion and exclusion criteria, information of 10,318 patients were extracted from the database. As shown in table 1, there were statistically significant differences among the baseline characteristics between patients those received chemotherapy versus those did not. Unbalanced characteristics were age group (p <0.001), year of diagnosis (p<0.001), tumor grade (p=0.001), size (p<0.001) and number of in-situ tumors (p<0.001). The five-year survival rate was 96.8 percent. After implementing the coarsened exact matching there were no significant differences in the covariates. The matched sample consisted of 3,222 patients with 1,611 in each arm (chemotherapy and no chemotherapy). Most patients were men (84.4%), White (97.6%) with solitary in-situ tumors (97.3%). More than a third had the tumor on the lateral wall (34%), grade IV tumors (35.6%) and sizes of 21-30 mm (33.3%).

**Table 1:**
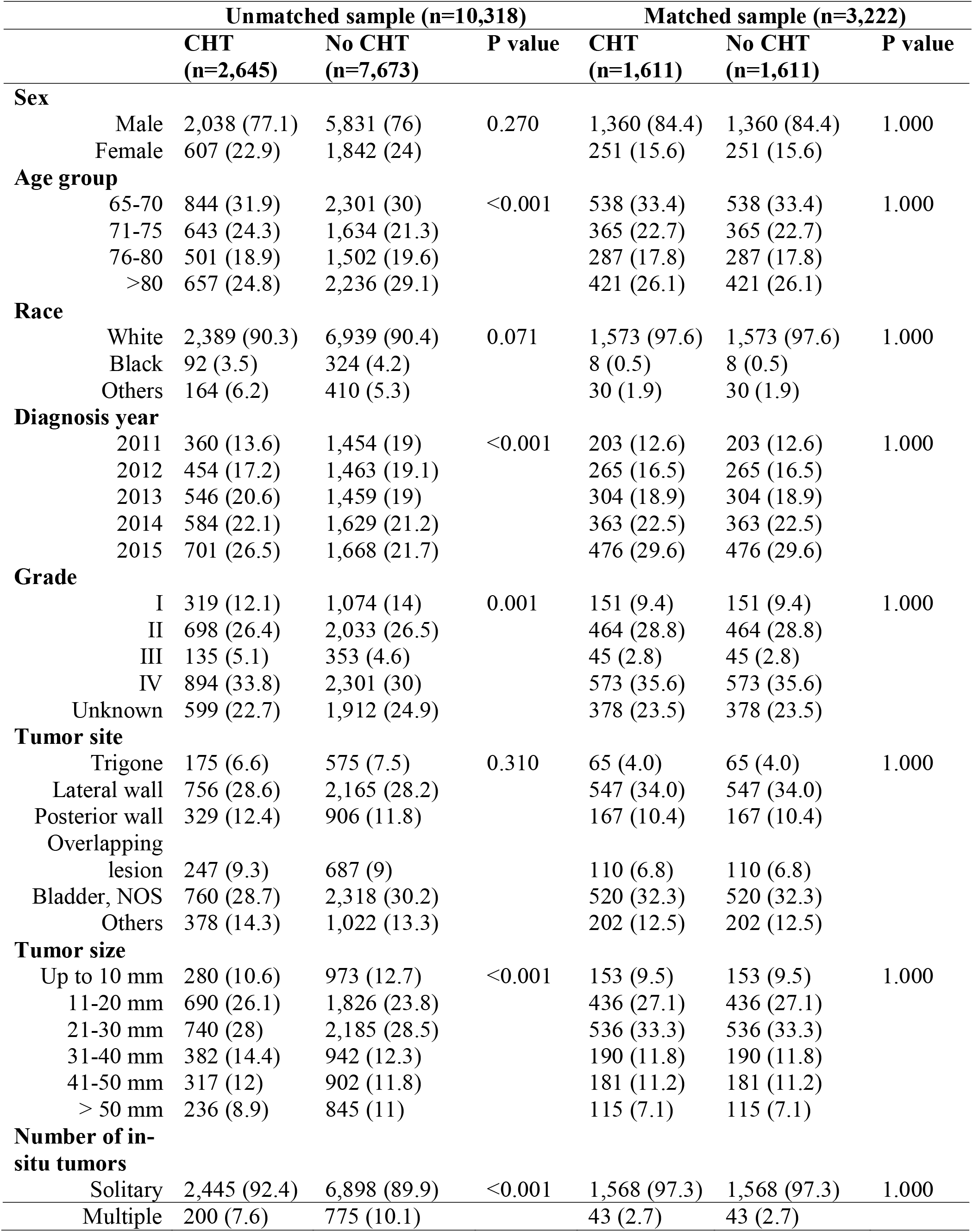
Clinical and pathological characteristics of patients by receipt of chemotherapy.

### Survival analysis

After matching and adjusting for covariates, multivariable Cox regression analyses (table 2) showed chemotherapy was associated with lower cancer-specific mortality (HR 0.63; 95% CI 0.42-0.94; p value 0.024). However, chemotherapy did not have any effect on overall mortality (HR 0.84; 95% CI 0.65-1.07; p value 0.159). Patients that are above 75 years and with multiple in-situ tumors had higher cancer specific and overall mortality. In addition, patients with grade IV tumors had higher cancer specific mortality. On the contrary, patients diagnosed in the year 2015 had significantly lower cancer-specific and overall mortality. Similar to the regression analyses, the Kaplan-Meier curves (Figure 1) show the protective effects of chemotherapy on cancer specific survival (p=0.032), but not on overall survival (p=0.34).

**Table 2:**
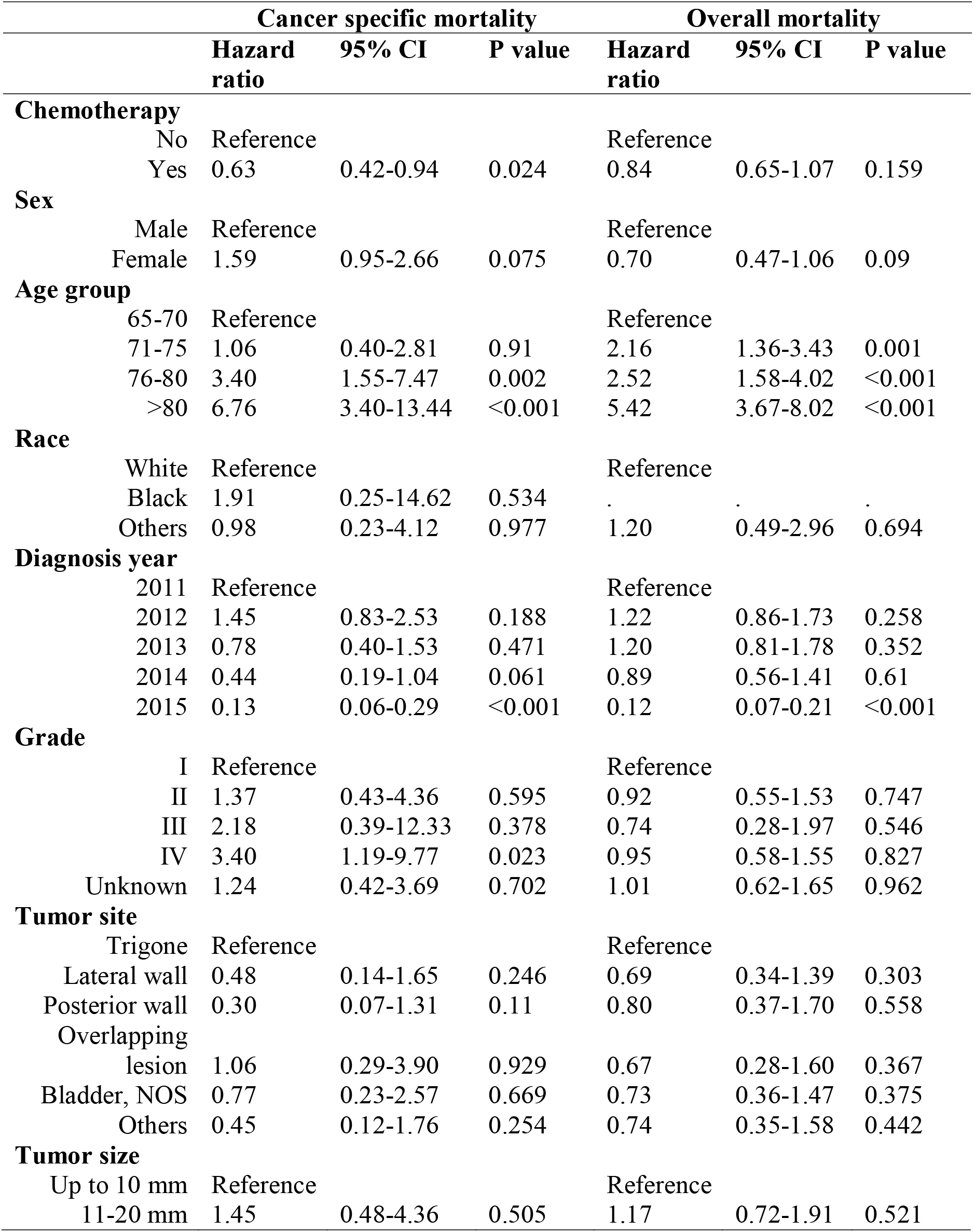
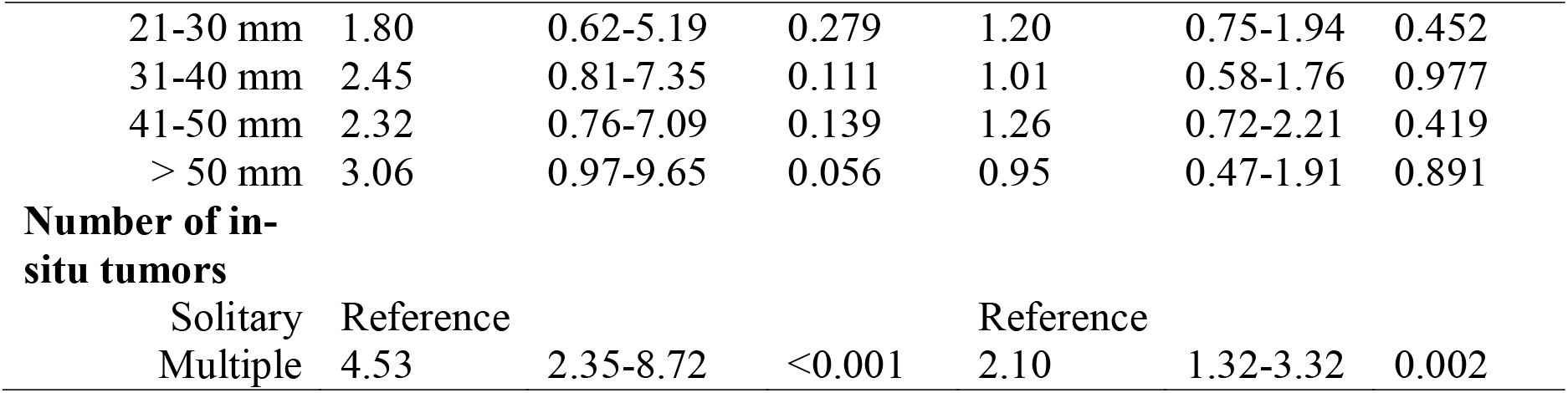
Cox regression models predicting cancer specific and overall mortality after matching (n=3,222)

|  |  | Cancer specific mortality |  |  | Overall mortality |  |  |
| --- | --- | --- | --- | --- | --- | --- | --- |
|  |  | Hazard ratio | 95% CI | P value | Hazard ratio | 95% CI | P value |
| **Chemotherapy** |  |  |  |  |  |  |  |
|  | No | Reference |  |  | Reference |  |  |
|  | Yes | 0.63 | 0.42-0.94 | 0.024 | 0.84 | 0.65-1.07 | 0.159 |
| **Sex** |  |  |  |  |  |  |  |
|  | Male | Reference |  |  | Reference |  |  |
|  | Female | 1.59 | 0.95-2.66 | 0.075 | 0.70 | 0.47-1.06 | 0.09 |
| **Age group** |  |  |  |  |  |  |  |
|  | 65-70 | Reference |  |  | Reference |  |  |
|  | 71-75 | 1.06 | 0.40-2.81 | 0.91 | 2.16 | 1.36-3.43 | 0.001 |
|  | 76-80 | 3.40 | 1.55-7.47 | 0.002 | 2.52 | 1.58-4.02 | <0.001 |
|  | >80 | 6.76 | 3.40-13.44 | <0.001 | 5.42 | 3.67-8.02 | <0.001 |
| **Race** |  |  |  |  |  |  |  |
|  | White | Reference |  |  | Reference |  |  |
|  | Black | 1.91 | 0.25-14.62 | 0.534 | . | . | . |
|  | Others | 0.98 | 0.23-4.12 | 0.977 | 1.20 | 0.49-2.96 | 0.694 |
| **Diagnosis year** |  |  |  |  |  |  |  |
|  | 2011 | Reference |  |  | Reference |  |  |
|  | 2012 | 1.45 | 0.83-2.53 | 0.188 | 1.22 | 0.86-1.73 | 0.258 |
|  | 2013 | 0.78 | 0.40-1.53 | 0.471 | 1.20 | 0.81-1.78 | 0.352 |
|  | 2014 | 0.44 | 0.19-1.04 | 0.061 | 0.89 | 0.56-1.41 | 0.61 |
|  | 2015 | 0.13 | 0.06-0.29 | <0.001 | 0.12 | 0.07-0.21 | <0.001 |
| **Grade** |  |  |  |  |  |  |  |
|  | I | Reference |  |  | Reference |  |  |
|  | II | 1.37 | 0.43-4.36 | 0.595 | 0.92 | 0.55-1.53 | 0.747 |
|  | III | 2.18 | 0.39-12.33 | 0.378 | 0.74 | 0.28-1.97 | 0.546 |
|  | IV | 3.40 | 1.19-9.77 | 0.023 | 0.95 | 0.58-1.55 | 0.827 |
|  | Unknown | 1.24 | 0.42-3.69 | 0.702 | 1.01 | 0.62-1.65 | 0.962 |
| **Tumor site** |  |  |  |  |  |  |  |
|  | Trigone | Reference |  |  | Reference |  |  |
|  | Lateral wall | 0.48 | 0.14-1.65 | 0.246 | 0.69 | 0.34-1.39 | 0.303 |
|  | Posterior wall | 0.30 | 0.07-1.31 | 0.11 | 0.80 | 0.37-1.70 | 0.558 |
|  | Overlapping lesion | 1.06 | 0.29-3.90 | 0.929 | 0.67 | 0.28-1.60 | 0.367 |
|  | Bladder, NOS | 0.77 | 0.23-2.57 | 0.669 | 0.73 | 0.36-1.47 | 0.375 |
|  | Others | 0.45 | 0.12-1.76 | 0.254 | 0.74 | 0.35-1.58 | 0.442 |
| **Tumor size** |  |  |  |  |  |  |  |
|  | Up to 10 mm | Reference |  |  | Reference |  |  |
|  | 11-20 mm | 1.45 | 0.48-4.36 | 0.505 | 1.17 | 0.72-1.91 | 0.521 |

|  |  |  |  |  |  |  |
| --- | --- | --- | --- | --- | --- | --- |
| 21-30 mm | 1.80 | 0.62-5.19 | 0.279 | 1.20 | 0.75-1.94 | 0.452 |
| 31-40 mm | 2.45 | 0.81-7.35 | 0.111 | 1.01 | 0.58-1.76 | 0.977 |
| 41-50 mm | 2.32 | 0.76-7.09 | 0.139 | 1.26 | 0.72-2.21 | 0.419 |
| > 50 mm | 3.06 | 0.97-9.65 | 0.056 | 0.95 | 0.47-1.91 | 0.891 |
| **Number of in-situ tumors** |  |  |  |  |  |  |
| Solitary | Reference |  |  | Reference |  |  |
| Multiple | 4.53 | 2.35-8.72 | <0.001 | 2.10 | 1.32-3.32 | 0.002 |

**Figure 1.**
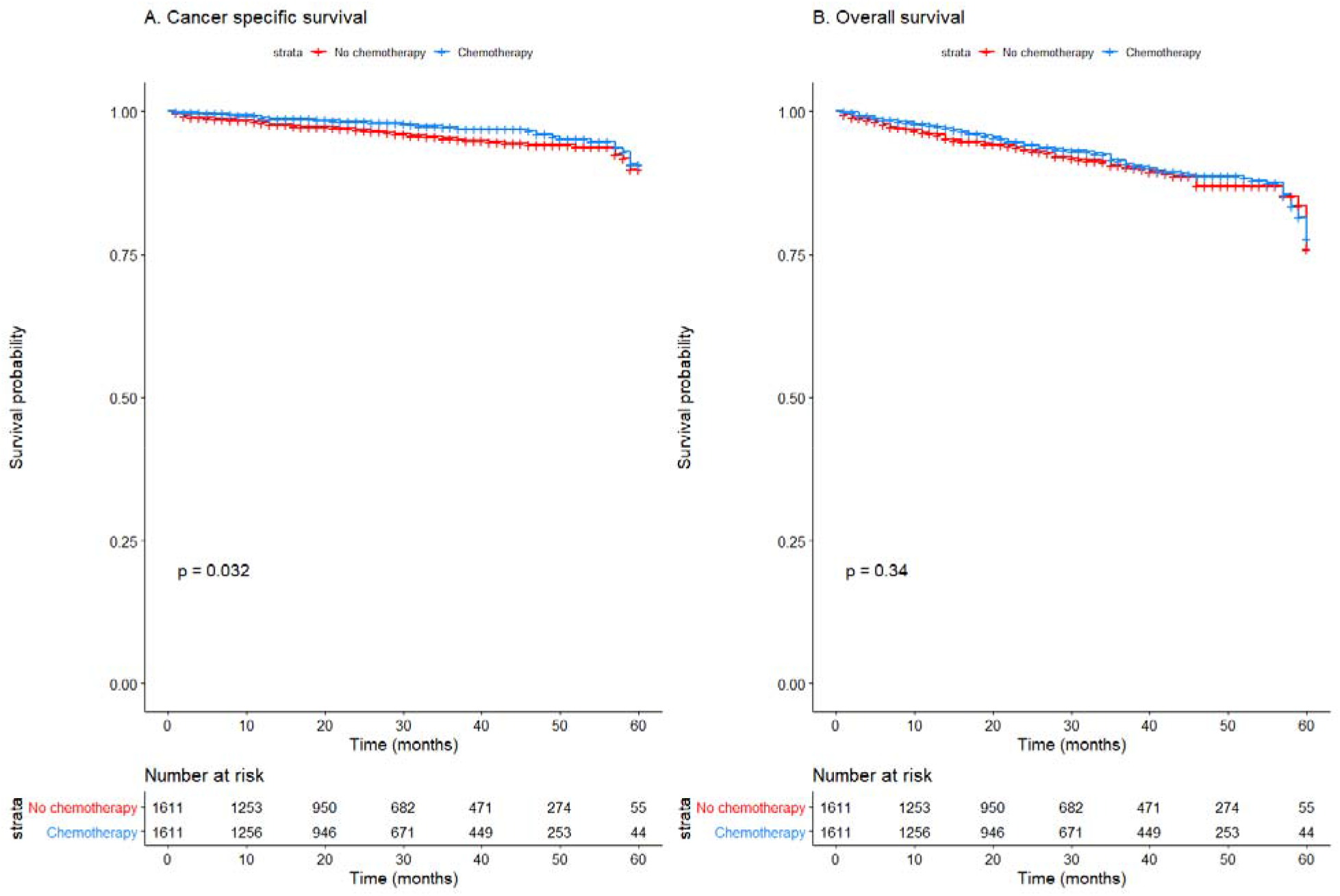
Effect of chemotherapy on cancer specific survival (A) and overall survival (B) in NMIBC patients undergoing TURBT.

## Discussion

In this study, we used the SEER database to retrospectively investigate the impact of additional chemotherapy on older patients with non-muscle invasive bladder cancer undergoing TURBT surgery. Chemotherapy improved cancer specific survival among the patients, but it had no effect on overall survival. To the best of our knowledge, this is the first large scale population-based real-world study to investigate the impact of chemotherapy on older patients with NMIBC undergoing TURBT surgery.

There could be two reasons for chemotherapy having no effect on overall survival. First, while meta-analyses prove the benefits of chemotherapy in NMIBC undergoing TURBT surgery,^6,11^ a few clinical trials have shown beneficial effects of chemotherapy on preventing recurrences of NMIBC in the short run (12 to 24 months).^12,13^ Beyond 24 months the protective effect of chemotherapy on recurrences were statistically not significant versus the control group. For instance, a trial by El-Ghobashy et. al. shows that the recurrence was 18.7% in the control group versus 3.2% in the chemotherapy group by 12 months.^13^ However, at 24 months it was 28.6% in control versus 26.9% in chemotherapy. Therefore, we could postulate that chemotherapy does not have any impact on long-term mortality in NMIBC patients. Secondly, overall survival could be influenced by non-cancer related co-morbid conditions.^14^ Our study findings on higher mortality among older patients, and those with grade IV and multiple in-situ tumors align with the current evidence.^15,16^ Literature shows that older patients are less likely to receive intravesical therapy.^16^ Contrary to some earlier studies, race was not significantly associated with mortality in our study.^17^ Lower mortality among the patients that were diagnosed in 2015 could reflect higher adherence to the current treatment guidelines on administering chemotherapy as well as recent advances in bladder cancer therapies.^18–20^

There are several limitations to our study. First, retrospective databases have the inherent selection bias even though we tried to control for confounders with matching. Second, SEER database does not contain the details of the chemotherapy regimens, their dosage nor the time of administration, e.g. adjuvant or neo-adjuvant. The database also does not have information on patients’ genetic constitution that might influence survival. Finally, our model did not include key socio-demographic features such as geographic location, household education and economic status, insurance as well as co-morbidities that might have an impact on the disease outcome. In conclusion, chemotherapy improved cancer specific survival among older patients with non-muscle invasive bladder cancer undergoing TURBT surgery, but it had no effect on overall survival. There is a need for more granular level real-world data on chemotherapy regimens and dosage to effectively investigate the effects of chemotherapy on survival of older patients with NMIBC that have undergone TURBT.

## Data Availability

SEER data publicly available

## Acknowledgements

We are grateful to the contributors of the Surveillance, Epidemiology, and End Results Program as well as to the National Cancer Institute for making this data publicly available.

